# Level of Cognitive Functioning among Elderly Patients in Urban area of Bangladesh: A Cross-Sectional Study

**DOI:** 10.1101/2024.08.22.24312384

**Authors:** Joynal Abedin Imran, Pradip Kumar Saha, Marzana Afrooj Ria, Syeda Saika Sarwar, Jannatul Ferdous Konok

## Abstract

Bangladesh is experiencing rapid urbanization and an increasing elderly population, particularly in urban areas. Cognitive decline is a common issue among elderly populations worldwide. This study aims to investigate the current state of cognitive decline among urban-dwelling older adults in Bangladesh to inform effective healthcare plans and programs. This cross-sectional study employed systematic random sampling among 150 participants at the outpatient department of NITOR in Dhaka, Bangladesh. Data were collected using a pretested structured standard questionnaire, and cognitive function was assessed with the Rowland Universal Dementia Assessment Scale (RUDAS). The study revealed a linear association between cognitive function and factors such as level of educational status, and history of eye sight problem. Conversely, age and diabetes showed an adverse correlation with cognitive function. Moreover, factors such as age, educational level and diabetes remained significant predictors of cognitive function in multivariate analysis. Cognitive impairment is significantly more common among the elderly in Bangladesh, with cognitive abilities being affected by a range of factors such as age, education level, diabetes, and vision issues.

## Introduction

Cognitive decline is a prevalent issue among elderly populations globally [1], and Bangladesh is not exempt from this trend. The nation, which is undergoing swift urbanization, is witnessing a rise in its elderly population, especially in urban regions [2]. The swift urbanization and the aging population in Bangladesh have heightened concerns about the mental health and cognitive abilities of the elderly. This demographic change presents substantial challenges as well as opportunities for healthcare systems.

Cognitive impairment, which can lead to dementia, is a primary cause of disability among the elderly and a leading factor in dependency for older adults across low-, middle-, and high-income countries [3]. This impairment is a significant public health issue as it greatly affects an individual’s quality of life, independence, and daily functioning. Mild cognitive impairment (MCI) represents an intermediate stage between normal cognitive function and dementia [4]. Research indicates that individuals with MCI are at a higher risk of developing dementia Currently, dementia impacts around 10 million people in the region, with its prevalence expected to double by 2030. Globally, approximately 35.6 million people suffer from dementia, a number projected to double to 65.7 million by 2030 and more than triple to 115.4 million by 2050. Dementia affects individuals worldwide, with over half (58%) residing in low- and middle-income countries; by 2050, this proportion is expected to exceed 70% In Bangladesh, Alzheimer’s and dementia-related deaths accounted for 14,993, or 2.09% of all deaths, according to the latest WHO data published in 2020 [5].

Currently, the majority of Bangladesh’s population is young; however, in the next 20 to 30 years, the proportion of older individuals is expected to increase significantly [6]. The urban elderly population in Bangladesh is growing rapidly, making it crucial to understand their cognitive functioning in order to develop effective healthcare strategies and interventions. There is a notable absence of government policies directly addressing the burden of cognitive impairment or dementia, largely due to the lack of reliable research that could inform policymakers, health professionals, and relevant authorities. Additionally, there is very limited research on cognitive impairment among the elderly in Bangladesh, which has not garnered sufficient attention from the research community. This study aims to address this gap by estimating the current prevalence of cognitive impairment and investigating its potential predictors among older adults in urban area of Bangladesh.

## Methodology

The research employed an observational method featuring quantitative data within a cross-sectional study design. Data were collected from elderly patients visiting the outpatient department at National Institute of Traumatology & Orthopaedic Rehabilitation (NITOR) in Dhaka during 3^rd^ March to 11^th^ June 2024. NITOR was chosen as the sampling area for its convenience. Inclusion criteria was set age from 60 years and above and people of either gender.

The following formula was used to fix the sample size:

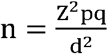

Here, Prevalence level, p = 0.109 or 10.9% [7]

Z = 1.96 for α = 95%

q = 1-p = (1-0.109) = 0.891

The desired sample size was 150. Informants were chosen using a systematic random sampling technique. The sampling interval was determined using the following formula:

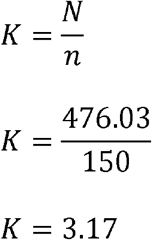

Here, *N* = Number of Population (173751 patients visited outpatient department, NITOR annually at 2022 [8] according to this data 476.03 patients visited daily), *n* = Number of sample size (150), *K* = Interval size

Following the sampling interval, every third elderly individual was approached and invited to participate in an interview. Prior to participation, each selected individual was required to provide written informed consent. In instances where a participant declined to participate, the next eligible individual, subsequent to every third person, was approached and invited to participate. The study was conducted in accordance with the declaration of Helsinki, and the protocol was approved by ethics committee of institutional review board of NITOR (Reference no. NITOR/PT/93/IRB/2024/4). Prior to commencing the study, participants were comprehensively apprised of the research objectives and methodologies. They were explicitly informed that their participation was entirely voluntary and that they could withdraw at any time. Furthermore, participants were assured that their data would be treated with the utmost confidentiality and anonymity, and that they would be fully informed of any potential risks and benefits associated with their involvement. Notably, no physical specimens were collected during the course of the study. Rather, researchers employed a standardized questionnaire to collect data, which is detailed below.

### Questionnaire

Elderly patients were interviewed using a structured questionnaire to gather information on sociodemographic characteristics and physical health status. Cognitive function was assessed using the Rowland Universal Dementia Assessment Scale (RUDAS), a standard tool developed in a multicultural context in Australia [9]. This brief, six-item test is designed for quick and simple administration, is portable, and is considered culturally and educationally unbiased. The RUDAS evaluates body orientation, praxis, drawing, judgment, memory, and language, and also assesses executive function impairment. Scores were calculated according to the RUDAS scoring manual. According to Basic [9], a score below 23 points indicates possible dementia, prompting referral for further neuropsychological evaluation. Scores above 27 points are considered normal, while scores between 23 and 27 points may suggest a mild neurocognitive disorder (MNCD). In our study, the survey instrument was translated into Bengali using forward and backward translation and field-tested among 30 elderly participants. Any inconsistencies of data were identified according to necessity and corrected during field level. Regarding content validity after translation process for RUDAS scale, Cronbach’s Alpha value was 0.9 and Cronbach’s Alpha based on standardized items was 0.906 indicated that the instrument has an acceptable level of validity.

### Statistical Analysis

The data analysis was performed using IBM SPSS version 26. Initially, descriptive statistical analysis was conducted to understand the participants’ characteristics, including their RUDAS scores. Pearson’s chi-square test or Fisher’s exact test, students t-test was utilized to explore the association between cognitive function and sociodemographic factors, physical health, and comorbidities. Bivariate correlations between cognitive function and continuous factors were assessed using correlation coefficients and categorical factors were assess using t-test and ANOVA to identify significant relationships. For multivariate analysis, a linear regression model was developed with the RUDAS score as the dependent variable. Only covariates significantly associated with cognitive function in the bivariate analyses were included in the multivariate model. This approach controlled for potential confounding effects and identified the independent effects of each factor on cognitive function. Statistical significance was set at a P-value of less than 0.05. This comprehensive statistical analysis approach allowed for an in-depth examination of the relationship between sociodemographic factors and cognitive function, controlling for confounding effects and identifying independent predictors of cognitive function in the elderly.

## Result

### Sociodemographic characteristics

In this study, 150 adults were enrolled, and none of the participants dropped out. The participants’ ages ranged from 60 to 85 years, with a mean age of 67.41 ± 6.31 years (95% confidence intervals = -1.76 to 2.08). The majority of participants were male (53.3%, n = 80), married (78.7%, n = 118), and had primary education (28%, n = 42) (Table 1). Regarding monthly family income, 68.6% (n = 103) were classified as low-income families. The mean age of males was slightly higher than that of females. Additionally, there were gender differences in marital status, education levels, and occupation.

**Table 1.** Sociodemographic characteristics of the participants (n = 150)

| Characteristics | All participants ($n = 150$) | Male ($n = 80$) | Female ($n = 70$) | Chi-square | *P* - value |
| --- | --- | --- | --- | --- | --- |
| Age (mean ± SD) | $67.41 \pm 6.31$ | $67.50 \pm 6.37$ | $67.31 \pm 6.30$ | - | 0.874<sup>a</sup> |
| Marital status (%) |  |  |  | 43.096<sup>b</sup> | <0.001 |
| Married | 118 (78.7) | 74 (92.5) | 44 (62.9) |  |  |
| Unmarried | 2 (1.3) | 1 (1.3) | 1 (1.4) |  |  |
| Widow/Widower | 30 (20) | 5 (6.3) | 25 (35.7) |  |  |
| Education (%) |  |  |  | 35.706<sup>c</sup> | <0.001 |
| No formal education | 17 (11.3) | 4 (5) | 13 (18.6) |  |  |
| Primary education (grade 1-5) | 42 (28) | 11 (13.8) | 31 (44.3) |  |  |
| Secondary education (grade 6-10) | 23 (15.3) | 13 (16.3) | 10 (14.7) |  |  |
| Higher secondary (grade 11-12) | 33 (22) | 22 (27.5) | 11 (15.7) |  |  |
| Graduation | 22 (14.7) | 19 (23.8) | 3 (4.3) |  |  |
| Post-graduation | 13 (8.7) | 11 (13.8) | 2 (2.9) |  |  |
| **Occupation (%)** |  |  |  | 148.568<sup>b</sup> | <0.001 |
| Retired | 32 (21.3) | 28 (35) | 4 (5.7) |  |  |
| Farmer | 11 (7.3) | 11 (13.8) | 0 |  |  |
| Businessman | 22 (14.7) | 22 (27.5) | 0 |  |  |
| Service holder | 12 (8) | 10 (12.5) | 2 (2.9) |  |  |
| Unemployed | 5 (3.3) | 5 (6.3) | 0 |  |  |
| Housewife | 61 (40.7) | 0 | 61 (87.1) |  |  |
| Others | 7 (4.7) | 4 (5) | 3 (4.3) |  |  |
| **Family monthly expense (Taka)<sup>d</sup>** |  |  |  | 3.886<sup>c</sup> | 0.274 |
| Below 20,000 | 29 (19.3) | 12 (15) | 17 (24.3) |  |  |
| 20,000 – 39,999 | 74 (49.3) | 39 (48.8) | 35 (50) |  |  |
| 40,000 – 59,999 | 35 (23.3) | 23 (28.7) | 12 (17.1) |  |  |
| 60,000 and above | 12 (8) | 6 (7.5) | 6 (8.6) |  |  |

<sup>a</sup>95% confidence intervals (CI) for mean difference = -1.76 to 2.08; <sup>b</sup>Fisher's exact correction test used for having 20% or more expected frequencies less than 5; <sup>c</sup>Pearson Chi-square; <sup>d</sup>1 US dollar = 109.39 Taka in February, 2024.

### Differences in characteristics of physical health and comorbidities related factors

Table 2 presents characteristics related to the physical health, smoking habits, sleep patterns, comorbidities, and cognitive function of the elderly population. The participants had a normal BMI, with a mean ± SD of 23.99 ± 4.06. The obesity rate was slightly higher in males (7.5%) compared to females (7.1%). Due to social norms in Bangladesh, none of the female participants smoked, while 15% of the male participants were current or former smokers. Most elderly individuals (54.7%) reported getting 6 to 7 hours of sleep per day. Diabetes, stroke, and cardiac disease were more prevalent among males, whereas sleeping difficulties, vision problems, arthritis, and asthma were more common among females.

**Table 2.**
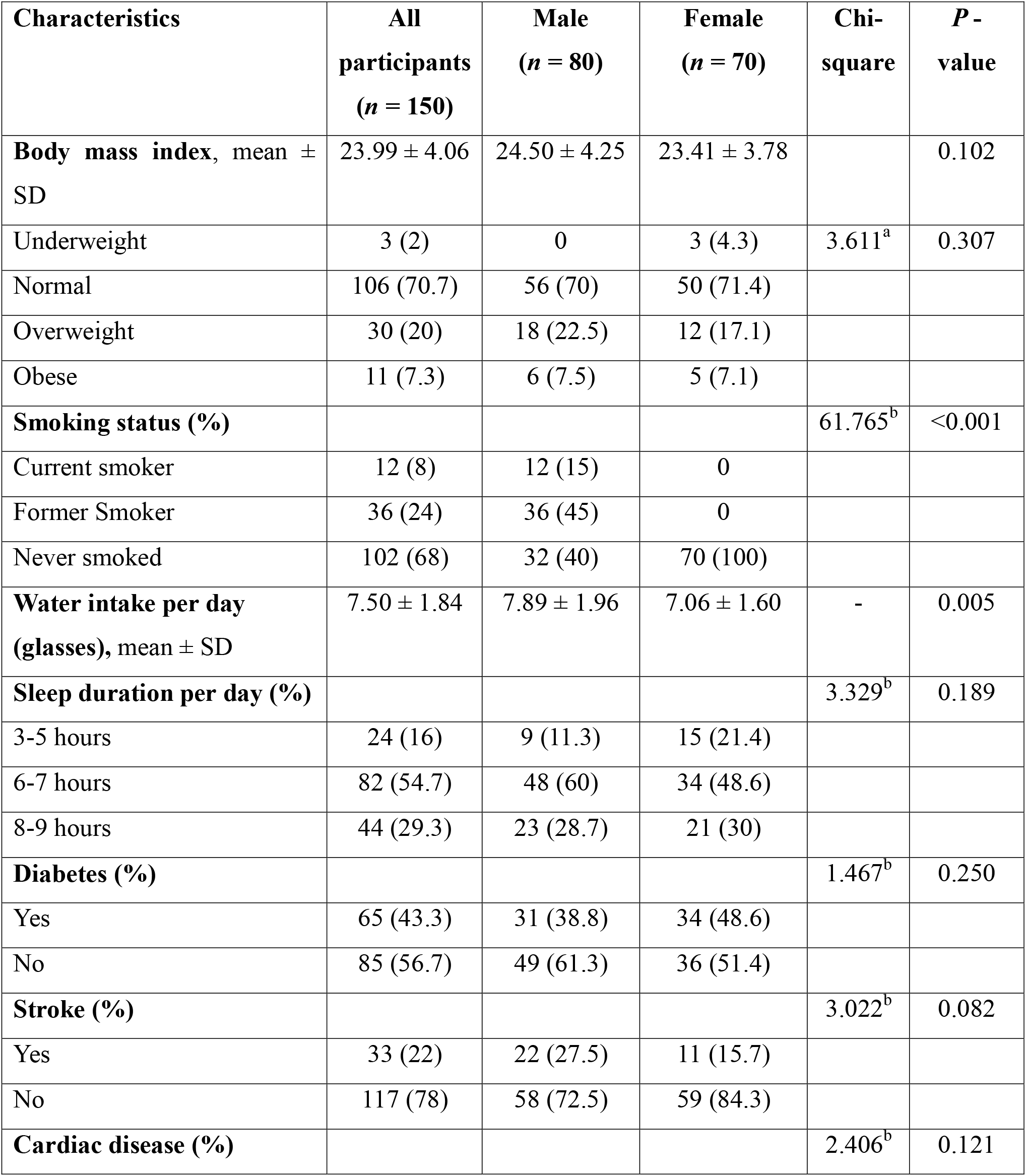
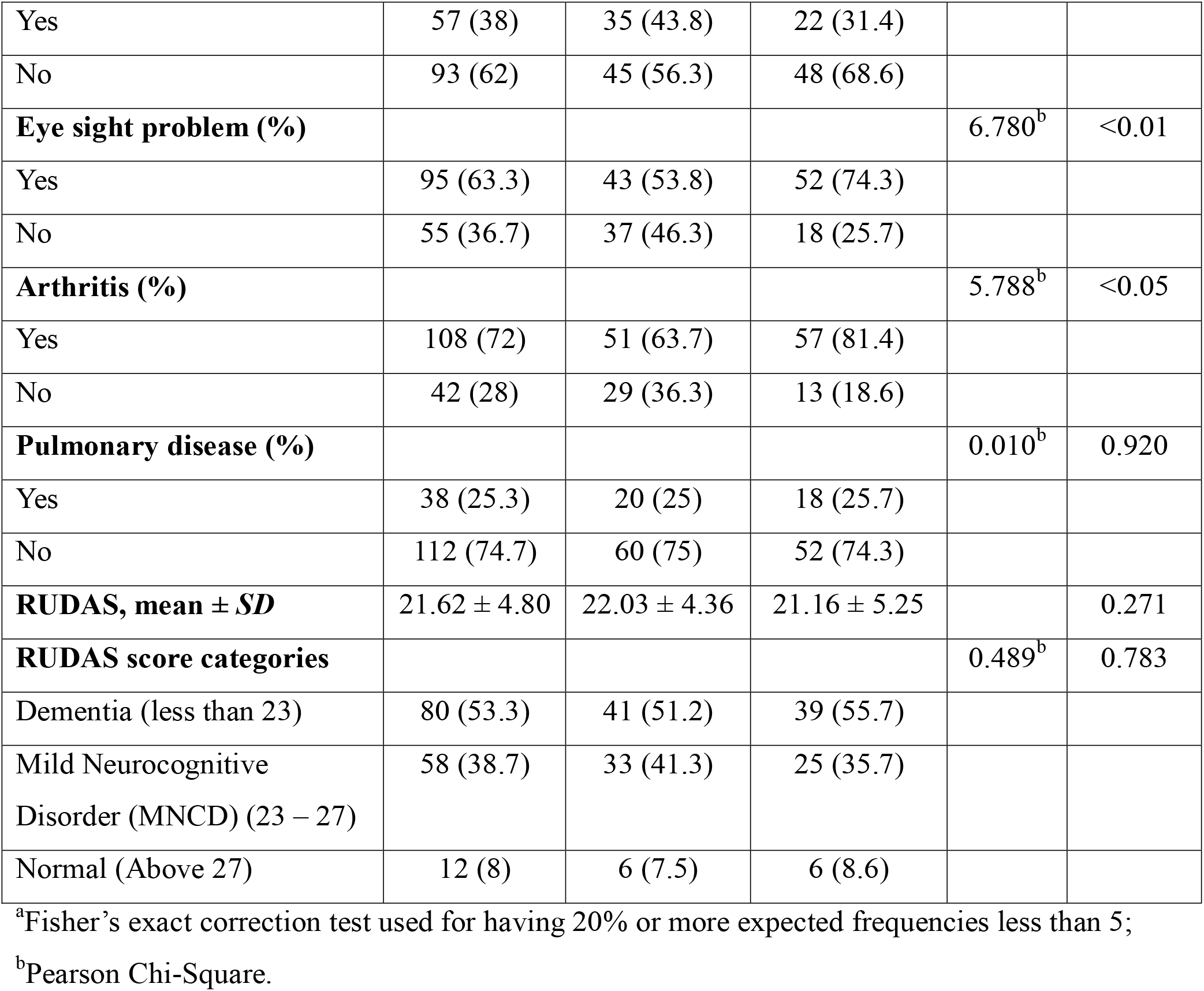
Characteristics of physical health, sleep hours, comorbidities and RUDAS score of the participants (n = 150)

#### RUDAS scores

The study participants (n = 150) generally exhibited poor cognitive function, with a mean ± SD score of 21.62 ± 4.80. Females had lower cognitive function scores compared to males (21.16 ± 5.25 vs. 22.03 ± 4.36). When categorizing cognitive function, the majority of participants (53.3%) were classified as having poor cognitive function (Dementia), over a quarter had mild neurocognitive disorder (MNCD), and only about 8% had normal cognitive function.

### Distribution of elderly people by Cognitive function score (RUDAS) categories

Among the 150 participants, the highest cognitive function score was 29 and the lowest was 8, out of a possible 30 points. The median cognitive function score was 22, with the 25th and 75th percentiles being 19 and 25, respectively. A large portion of the participants (53.3%) were identified as having dementia, while only 8% were observed to have normal cognitive function (Figure 1).

**Figure 1.**
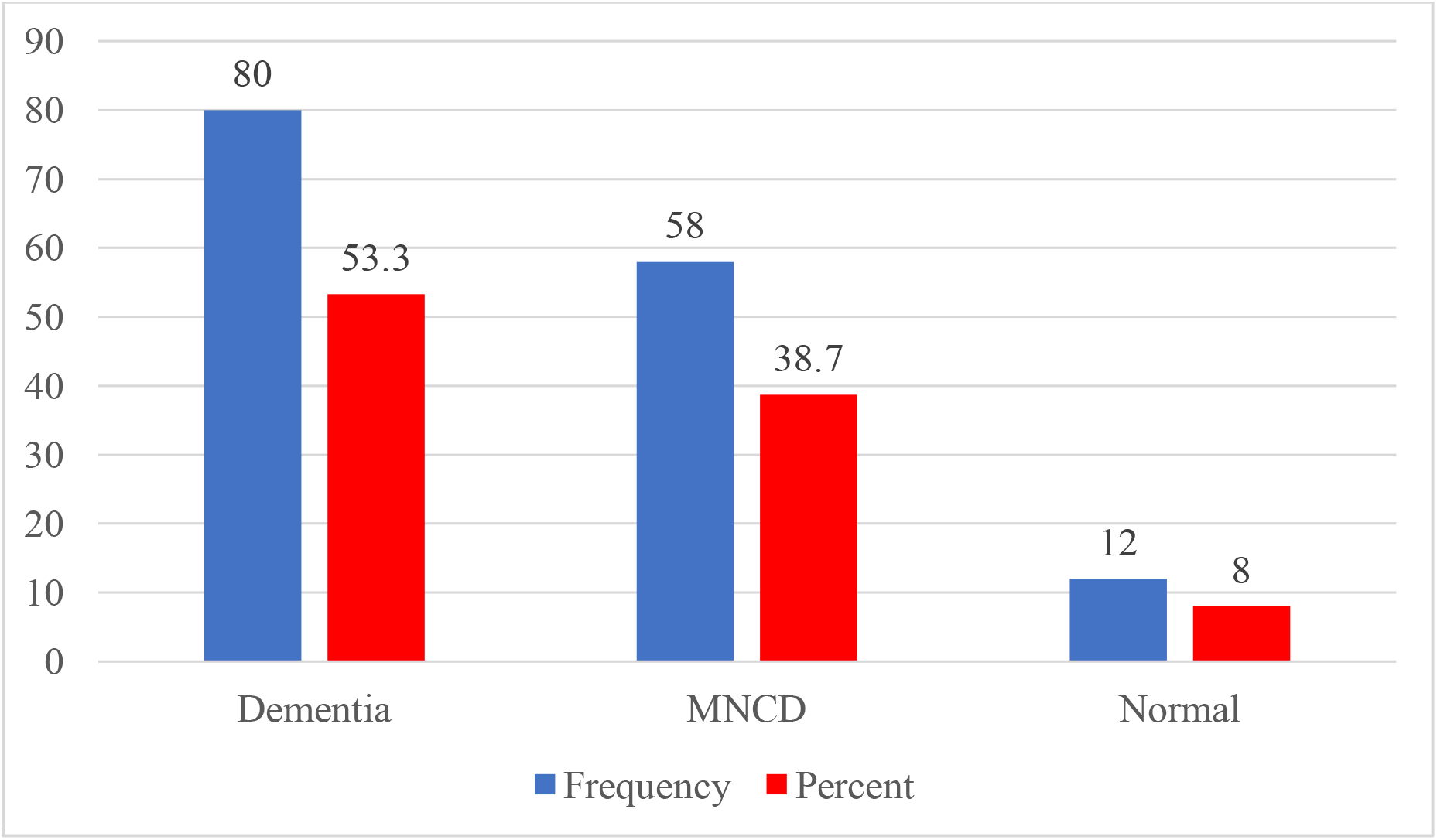
Distribution of the participants by RUDAS scores (n = 150)

### Correlation between Sociodemographic Factors and Cognitive function (RUDAS) Scores

A bivariate correlation test (Table 3) revealed that cognitive function was inversely correlated with age (r = -0.451, P = 0.000), indicating poorer cognitive function with increasing age among the elderly population.

**Table 3.** Correlation matrix between RUDAS scores with sociodemographic and comorbidities factors in elderly population (n = 150)

|  |  | Age | Monthly<br>family<br>expense | BMI | Water<br>intake | Sleep<br>duration | RUDAS |
| --- | --- | --- | --- | --- | --- | --- | --- |
| Age | *r* | 1 | 0.073 | 0.078 | -0.033 | 0.085 | -0.451** |
|  | *P* |  | 0.375 | 0.341 | 0.690 | 0.303 | 0.000 |
| Monthly<br>family<br>expense | *r* |  | 1 | 0.094 | -0.049 | -0.062 | 0.109 |
|  | *P* |  |  | 0.251 | 0.548 | 0.453 | 0.184 |
| BMI | *r* |  |  | 1 | -0.080 | 0.163* | 0.138 |
|  | *P* |  |  |  | 0.331 | 0.047 | 0.092 |
| Water intake | *r* |  |  |  | 1 | 0.020 | 0.104 |
|  | *P* |  |  |  |  | 0.806 | 0.207 |
| Sleep duration | *r* |  |  |  |  | 1 | 0.039 |
|  | *P* |  |  |  |  |  | 0.637 |

\*\*Correlation is significant at the 0.01 level (2-tailed); \*Correlation is significant at the 0.05 level (2-tailed)

A bivariate t-test and ANOVA test that revealed Education (P = 0.009), Diabetes (P = 0.038), and Eye Sight Problems (P = 0.009) are significantly associated with RUDAS Scores implying they were significant predictors understanding cognitive performance. (Table 4)

**Table 4.** Association between RUDAS scores with sociodemographic and comorbidities factors in elderly population (n = 150)

| Variables | RUDAS Score |  | *P*-value |
| --- | --- | --- | --- |
|  | Mean | SD |  |
| **Gender** |  |  | 0.271<sup>a</sup> |
| Male | 22.03 | 4.36 |  |
| Female | 21.16 | 5.25 |  |
| **Marital Status** |  |  | 0.528<sup>b</sup> |
| Married | 21.85 | 4.58 |  |
| Unmarried | 21.50 | 6.36 |  |
| Widow/widower | 20.73 | 5.59 |  |
| **Education** |  |  | 0.009<sup>a</sup> |
| Graduation & below | 21.34 | 4.81 |  |
| Post graduation | 24.62 | 3.66 |  |
| **Occupation** |  |  | 0.347<sup>b</sup> |
| Retired | 22.03 | 4.43 |  |
| Farmer | 20.36 | 5.03 |  |
| Businessman | 22.23 | 3.68 |  |
| Service holder | 24.08 | 3.50 |  |
| Unemployed | 19.20 | 5.81 |  |
| Housewife | 21.05 | 5.36 |  |
| Others | 22.29 | 4.96 |  |
| **Smoking status** |  |  | 0.636<sup>b</sup> |
| Current smoker | 21.83 | 3.79 |  |
| Former smoker | 22.25 | 4.14 |  |
| Never smoked | 21.37 | 5.13 |  |
| **Diabetes** |  |  | 0.038<sup>a</sup> |
| Present | 20.68 | 5.04 |  |
| Absent | 22.34 | 4.52 |  |
| **Stroke** |  |  | 0.446<sup>a</sup> |
| Present | 20.97 | 5.74 |  |
| Absent | 21.80 | 4.52 |  |
| **Cardiac disease** |  |  | 0.606<sup>a</sup> |
| Present | 21.37 | 4.44 |  |
| Absent | 21.77 | 5.04 |  |
| **Eye sight problem** |  |  | 0.009<sup>a</sup> |
| Present | 20.84 | 5.16 |  |
| Absent | 22.96 | 3.81 |  |
| **Arthritis** |  |  | 0.292<sup>a</sup> |
| Present | 21.37 | 4.91 |  |
| Absent | 22.26 | 4.51 |  |
| **Pulmonary disease** |  |  | 0.919<sup>a</sup> |
| Present | 21.68 | 4.32 |  |
| Absent | 21.60 | 4.98 |  |

<sup>a</sup>t-test; <sup>b</sup>ANOVA

### Linear Regression Analysis to Predict Cognitive Function

The variables that were statistically significant in the bivariate analysis were included in the regression model. The dependent variable was the continuous cognitive function (RUDAS) score. The selected independent variables were: age, educational status (0 = graduation & below, 1 = post-graduation), diabetes (0 = yes, 1 = no), and eyesight problems (0 = yes, 1 = no). Before developing the model multicollinearity and residual assumptions were analyzed. No multicollinearity was observed among the independent variables. The standard residual value was minimum -2.896 and maximum 2.357 indicating no outliers in the dependent variable. The Durbin-Watson test value was 1.276 indicating data points were independent. To analyze the normality of the residuals and constant variance; histogram, normal probability (P-P) plot and a scatter plot of the residuals were produced. The distribution of the residuals was seen normal in the histogram and P-P plot. The variance (homoscedasticity) was constant in the scatter plot.

Table 5 data shows that the statistically significant predictors of cognitive function including: age (*P* = <0.001), educational status (*P* = 0.046), history of diabetes (*P* = 0.005). Eye sight problem was excluded from the model because of P > 0.05.

**Table 5.** Factors predicting cognitive function in elderly people (n= 150)

| Variable | β- coefficient | SE | Standardized β-coefficient | P-value | 95% CI for $\beta$ |
| --- | --- | --- | --- | --- | --- |
| Constant | 41.96 | 3.74 |  | <0.001 |  |
| Age | -0.33 | 0.06 | -0.43 | <0.001 | -0.43 to -0.22 |
| Educational status | 2.4 | 1.20 | 0.14 | 0.046 | 0.05 to 4.79 |
| Diabetes | 1.95 | 0.68 | 0.20 | 0.005 | 0.60 to 3.30 |

Variables excluded from the model: Eye sight problem ( $P = 0.058$ )

The predicted model for the data was as follows:

Cognitive function (RUDAS) = 41.96 – 0.33 (age) + 2.4 (educational status) + 1.95 (history of diabetes)

## Discussion

Findings based on the RUDAS scale in this study indicate that a significant number of participants aged 60 years and older exhibited mild neurocognitive impairment and dementia. Similar results have been reported in several studies, which found that cognitive performance significantly decreases with increasing age [10-12]. Additionally, our study found that females had more impaired cognitive function than males, a trend also noted by Beam et al. and Oishee et al. [11,13].

In this study, educational qualifications of the elderly showed a significant relationship with cognitive function. Higher educational status was associated with better cognitive function in the elderly. Research supports that higher education levels correlate with improved cognitive health and can serve as a protective factor against cognitive decline [14,15]. Additionally, the level of education moderates the relationship between depressive symptoms and cognitive functioning, with lower education levels being linked to poor cognitive outcomes [16].

This study found a higher prevalence of diabetes among female participants, which was significantly associated with cognitive decline. Diabetic elderly participants had lower cognitive function compared to non-diabetic participants. Elderly individuals with diabetes are at increased risk for cognitive dysfunction and dementia, affecting their overall cognitive functioning. Research supports that diabetes, whether diagnosed or undiagnosed, is linked to poor cognitive function, particularly in the elderly [17,18]. Cognitive impairment among those aged 65 and older, as measured by the Mini-Mental State Examination (MMSE), was significantly higher among diabetic patients [19].

Although this study had found no significant relationship between eyesight problems and cognitive function but some study supports a strong association between cognitive function and visual ability in the elderly. Studies have shown that individuals with mild cognitive impairment (MCI) have lower functional visual acuity compared to those with normal cognition [20], and worsening vision is linked to declining cognitive function over time [21]. Improved cognitive function is associated with better best-corrected visual acuity, highlighting the importance of addressing visual impairments to mitigate cognitive decline in older adults [22].

### Limitations

This study was conducted in a specific area of Bangladesh, chosen purposively due to the cooperation of local authorities. To address these limitations, moving forward, future studies should explore additional factors that may influence cognitive function and incorporate longitudinal designs to better understand the trajectory of cognitive decline over time as this study provided a snapshot of an elderly population at a specific moment, making it difficult to assess temporal relationships or trends.

## Conclusions

Efforts to develop tailored interventions and policies should consider the multifaceted nature of cognitive health and the diverse needs of aging populations in Bangladesh and beyond. By addressing these predictors and implementing targeted interventions, Government & NGO’s can work towards improving cognitive function and quality of life for elderly individuals in community level at Bangladesh.

## Supporting information

Supplement: Informed Consent form

Supplement: Questionnaire

## Data Availability

All relevant data are within the manuscript and its Supporting Information files

## Supporting information

S1 Appendix. Informed consent form. The included written consent form administered to the participants.

S2 Appendix. Questionnaire. The included questions comprised the questionnaire administered to the participants.

S3 Appendix. The included data of the participants.

## Acknowledgement

We would like to recognize the efforts of Sanjib Saha, Associate researcher, Lund University, Sweden who helped us through this study.

## Author contributions

Conceptualization: Joynal Abedin Imran and Pradip Kumar Saha

Methodology: Joynal Abedin Imran;

Software: Joynal Abedin Imran

Validation: Pradip Kumar Saha Formal Analysis: Joynal Abedin Imran

Investigation: Marzana Afrooj Ria and Syeda Saika Sarwar

Resources: Marzana Afrooj Ria and Jannatul Ferdous Konok

Data Curation: Syeda Saika Sarwar

Writing – Original Draft Preparation: Joynal Abedin Imran

Writing – Review & Editing: Joynal Abedin Imran, Marzana Afrooj Ria and Jannatul Ferdous Konok

Visualization: Joynal Abedin Imran and Pradip Kumar Saha

Supervision: Pradip Kumar Saha

Project Administration: Pradip Kumar Saha

### Funding

No specific funding

