## Supplement: Informed Consent form for "Level of Cognitive Functioning among Elderly Patients in Urban area of Bangladesh: A Cross-Sectional Study"

My name is (mention the interviewer’s name).

**Project Goal**:

Congratulations! You have been selected to take part in this research project. This study aims to know the level of cognitive functioning among elderly people, like you.

**Procedures**: I would like to interview you if you want to take part in this study. This interview may take about 15 minutes. For this study, we are not taking any blood, urine, or other samples from you. I will explain a few other things about your confidentiality, rights, and any risks and benefits if you choose to participate in this study. I assure you that you will not lose any benefits that you normally get from this hospital even if you don’t participate in this study.

**Confidentiality:** All the information given by you in this study will be kept confidential and will be used only for academic and research purposes.

**Participant's rights, risks, and benefits:** Please understand that your participation in this study is completely voluntary and that you can withdraw from the study at any time once enrolled. Once enrolled in this study, you will participate in answering some questions. However, you have the right to decline to answer any questions during the interview. Also understand that although there is no direct participant in participating in the study, the results of this study will help in the future for a better understanding of the quality of life and health needs of elderly people.

**For any further questions, you can reach one of us by calling the above phone numbers.**

**Participant's consent:** I have been informed well about this study. I give my consent consciously and voluntarily to take part in this study.

______________________	__________________

**Signature of the Participant	Date**

**Mobile No.: ___________________**

**_______________________	___________________**

**Signature of researcher**	**Date**
