## Supplement: Questionnaire for "Level of Cognitive Functioning among Elderly Patients in Urban area of Bangladesh: A Cross-Sectional Study"

**SUPPORTING INFORMATION 2 -APPENDIX: Questionnaire**

**Participant ID No:**	/__/__/	Date:

**Name of the participant:**

**Address:**

**Section 1- Socio demographic factor:**

| **Serial No.** | **Question** | **Response** | **Code No.** |
| --- | --- | --- | --- |
| 1 | What is your age? | /__/__/__/__/__/__/ Years |  |
| 2 | Gender | - Male - Female | 0  1 |
| 3 | What is your marital status? | - Married - Unmarried - Widow - Widower - Divorced | 0  1  2  3  4 |
| 4 | What is your educational qualification? | - No formal education - Primary education - Secondary education - Higher secondary - Graduation - Post-graduation | 0  1  2  3  4  5 |
| 5 | What is your occupation? | - Retired - Farmer - Businessman - Service holder - Unemployed - House wife - Others | 0  1  2  3  4  5  6 |
| 6 | If not retired or unemployed, monthly income from your occupation? | BDT |  |
| 7 | What is your monthly family expense? | BDT |  |

**Section 2-** **Physical health related factors:**

| **Serial No.** | **Question** | **Response** | **Code No.** |
| --- | --- | --- | --- |
| 8 | Height | Inch |  |
| 9 | Weight | kg |  |
| 10 | BMI | Kg/m^2^ |  |
| 11 | How many glasses of water do you drink every day? | Times |  |
| 12 | Your smoking status? | - Current smoker - Former smoker - Never smoked | 0  1  2 |
| 13 | How much do you sleep per day? | Hour |  |

**Section 3- Comorbidities related factors**

| **Serial No.** | **Question** | **Response** | **Code No.** |
| --- | --- | --- | --- |
| 14 | Do you have any history of diabetes? | - Yes - No | 0  1 |
| 15 | If yes, how many years? |  |  |
| 16 | Do you have any history of stroke? | - Yes - No | 0  1 |
| 17 | Do you have any history of cardiac disease? | - Yes - No | 0  1 |
| 18 | Do you have any history of eye sight problem? | - Yes - No | 0  1 |
| 19 | Do you have any history of arthritis? | - Yes - No | 0  1 |
| 20 | Do you have any history of pulmonary disease? | - Yes - No | 0  1 |

**Section 4- Rowland Universal Dementia Assessment Scale (RUDAS)**

**Date: ___/___/___	Patient Name:________________________**

| **Item** |  | **Max Score** |
| --- | --- | --- |
| **Memory**  1. (Instruction) I want you to imagine that we are going shopping. Here is a list of grocery items. I would like you to remember the following items which we need to get from the shop. When we get to the shop in about 5 mins. time I will ask you what it is that we have to buy. You must remember the list for me.  **Tea, Cooking Oil, Eggs, Soap** Please repeat this list for me (ask person to repeat the list 3 times). (If person did not repeat all four words, repeat the list until the person has learned them and can repeat them, or, up to a maximum of five times.) |  |  |
| **Visuospatial Orientation** 2. I am going to ask you to identify/show me different parts of the body. (*Correct = 1)*. Once the person correctly answers 5 parts of this question, do not continue as the maximum score is 5.   (1) Show me your right foot (2) Show me your left hand (3) With your right hand touch your left shoulder (4) With your left hand touch your right ear (5) Which is (indicate/point to) my left knee (6) Which is (indicate/point to) my right elbow (7) With your right hand indicate/point to my left eye (8) With your left hand indicate/point to my left foot |  |  |
| **Praxis**  3. I am going to show you an action/exercise with my hands. I want you to watch me and copy what I do.  Copy me when I do this. . . (One hand in fist, the other palm down on table – alternate simultaneously.) Now do it with me: Now I would like you to keep doing this action at this pace until I tell you to stop- approximately 10 seconds (Demonstrate at moderate walking pace).  Score as:  *Normal = 2 (very few if any errors; self-corrected, progressively better; goo maintenance; only very slight lack of synchrony between hands)*  *Partially Adequate = 1 (noticeable errors with some attempt to self-correct; some attempt at maintenance; poor synchrony)*  *Failed = 0 (cannot do the task; no maintenance; no attempt whatsoever)* |  |  |
| **Visuoconstructional Drawing**  4. Please draw this picture exactly as it looks to you (show cube on back of page). *(Yes=1)*  Score as:   (1) Has person drawn a picture based on a square? (2) Do all internal lines appear in person’s drawing  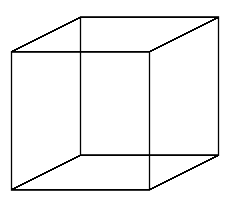  (3) Do all external lines appear in person’s drawing?  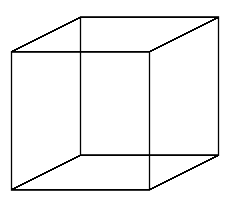 Judgement:  5. You are standing on the side of a busy street. There is no pedestrian crossing and no traffic lights. Tell me what you would do to get across to the other side of the road **safely**. (If person gives incomplete response that does not address both parts of answer, use prompt: “Is there anything else you would do?”) Record exactly what patient says and circle all parts of response which were prompted.  Score as:  Did person indicate that they would look for traffic? *(YES = 2; YES PROMPTED = 1; NO = 0)*  Did person make any additional safety proposal? *(YES =2; YES PROMPTED =1; NO = 0)* |  |  |
| **Memory recall** 1. (Recall) We have just arrived at the shop. Can you remember the list of groceries we need to buy? (Prompt: If person cannot recall any of the list, say “The first one was ‘tea’.” *(Score 2 points each for any item recalled which was not prompted – use only ‘tea’ as a prompt.)* *Tea* *Cooking Oil* *Eggs* *Soap* **Language** 6. I am going to time you for one minute. In that one minute, I would like you to tell me the names of as many different animals as you can. We’ll see how many different animals you can name in one minute. (Repeat instruction if necessary). Maximum score for this item is 8. If person names 8 new animals in less than one minute there is no need to continue. 1. ……………………………. \| 5. …………………………….; 2. ……………………………. \| 6. …………………………….; 3. ……………………………. \| 7. …………………………….; 4. ……………………………. \| 8. ……………………………. |  |  |
|  |  | **…/8** |
| **TOTAL SCORE =** |  | **/30** |

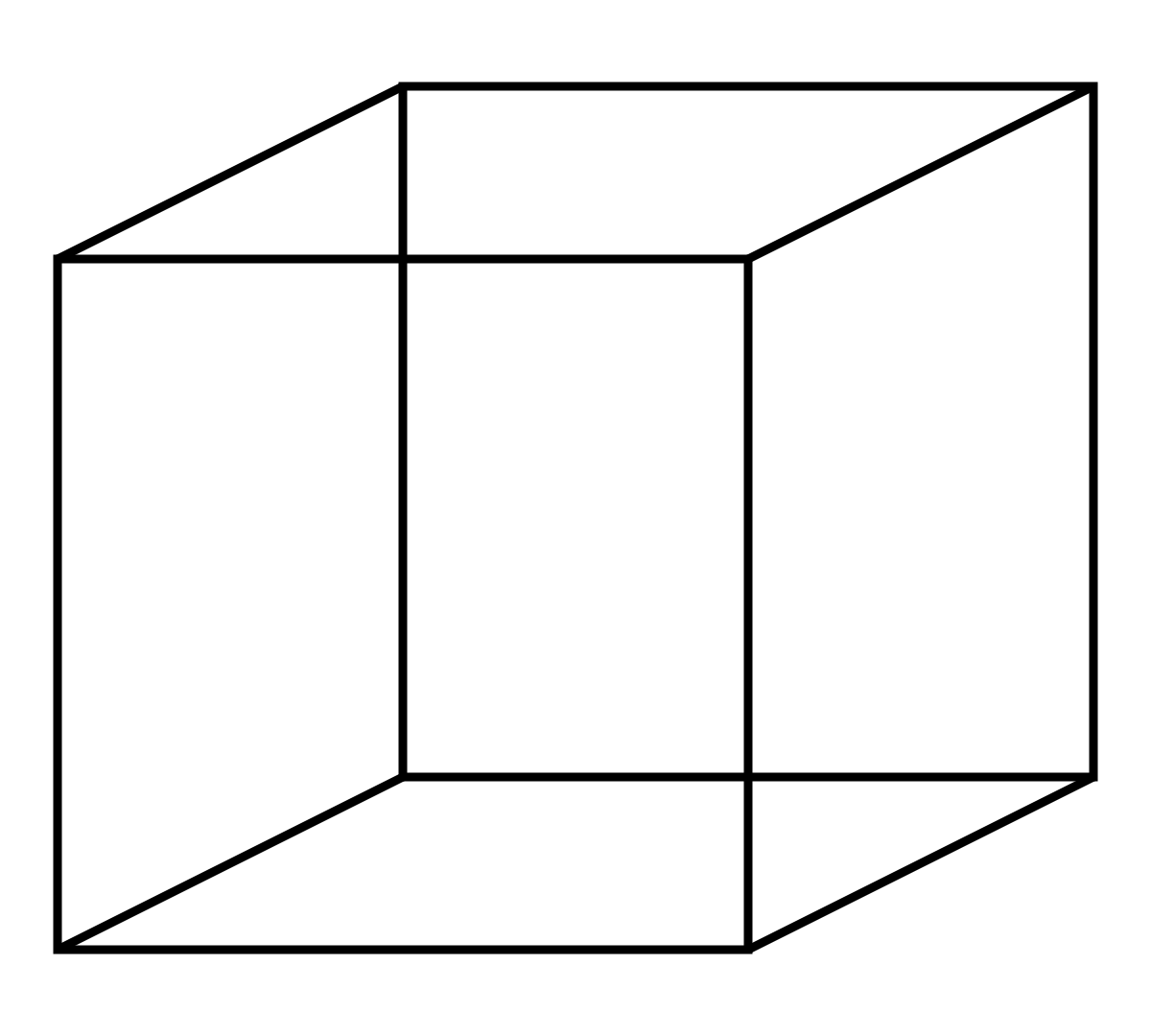
